# Large-scale characterization of gender differences in diagnosis prevalence and time to diagnosis

**DOI:** 10.1101/2023.10.12.23296976

**Authors:** Tony Yue Sun, Jill Hardin, Harry Reyes Nieva, Karthik Natarajan, Ru-fong Cheng, Patrick Ryan, Noémie Elhadad

## Abstract

We carry out an analysis of gender differences in patterns of disease diagnosis across four large observational health datasets and find that women are routinely older when first assigned most diagnoses. Among 112 acute and chronic diseases, women experience longer lengths of time between symptom onset and disease diagnosis than men for most diseases regardless of metric used, even when only symptoms common to both genders are considered. These findings are consistent for patients with private as well as government insurance. Our analysis highlights systematic gender differences in patterns of disease diagnosis and suggests that symptoms of disease are measured or weighed differently for women and men. Data and code leverage the open-source common data model and analytic code and results are publicly available.

**One-Sentence Summary:** In large populations, across insurance coverage and many conditions, women are older than men when diagnosed and experience longer time to diagnosis.

## Main Text

One of the major tenets of clinical medicine is the prompt diagnosis of disease. Armed with a timely diagnosis, clinicians can intervene early, potentially curb the progression of disease, and in turn prevent long-term health consequences. For patients, obtaining a diagnosis can reduce the anxiety and distress related to experiencing unacknowledged or undiagnosed symptomatic disease. Because the effective diagnosis of a condition depends on recognizing its signs and symptoms, accounting for differences in symptom presentations among groups of individuals is critical to ensure equitable and timely diagnosis. Other factors, such as access to care, systemic racism, and gender bias, can also impact timely receipt of a diagnosis. In this work, we interrogate differences in diagnosis patterns between women and men.

A nascent paradigm in clinical research seeks to identify the role of sex and gender^1^ in medicine in an effort to improve healthcare outcomes for all (*2*, *3*). A large body of research has documented sex-differences and gender-based disparities across clinical medicine, epidemiology, pathophysiology, clinical manifestation, and outcome management (*4*). The most often-cited examples are in cardiology, where literature shows that women and men experience different symptoms for acute myocardial infarction. Women are less likely to present with chest pain—the main presenting symptom for men— but instead report higher rates of fatigue, dyspnea, nausea, and other forms of pain (*5–7*). Because healthcare providers are traditionally trained using diagnostic criteria based on presentation in men, they can fail to recognize heart attacks in women, which ultimately contributes to decreased survival outcomes for women (*8–11*). With these findings in mind, the research community has pushed toward more equal representation in clinical science. For example, the National Institutes of Health now require new clinical trials contain adequate data from both sexes (*12*). Despite the importance of this issue and more recent body of research associated with it, significant gaps in knowledge persist; for the vast majority of outstanding conditions and diseases, gendered differences in disease prevalence, age of onset, presentation, and treatment remain unknown (*13*).

In this study, we systematically characterize differences in patterns of diagnosis among women and men for a broad spectrum of diseases and conditions. Our analyses leverage more than 200 million observational longitudinal records in the United States and span privately-insured claims data, government program claims data, and the electronic health record (EHR) data of a large urban medical center. Specifically, we seek to assess gender differences in diagnosis by answering two research questions:

1. Across all diagnosed conditions in a population, are there differences in condition prevalence, risk ratio, increased risk, and age of onset between women and men? To address this question, we carry out a **population-wide characterization** analysis.
2. Across cohorts of patients with the same diagnosis, are there differences in the time to diagnosis or diagnostic delay between women and men? Because we leverage disease-specific sets of condition criteria (i.e., phenotypes) to identify such cohorts when addressing this question, we refer to our second analysis as **phenotype-specific characterization**.

## Data and analytical strategy

We perform our analyses on four datasets that cumulatively comprise the de-identified longitudinal records of over 200 million Americans. **Fig. 1** depicts the analyses carried out on each dataset separately. Code for generating phenotype definitions and performing analyses are publicly available on GitHub (*14*).

**Fig. 1.**
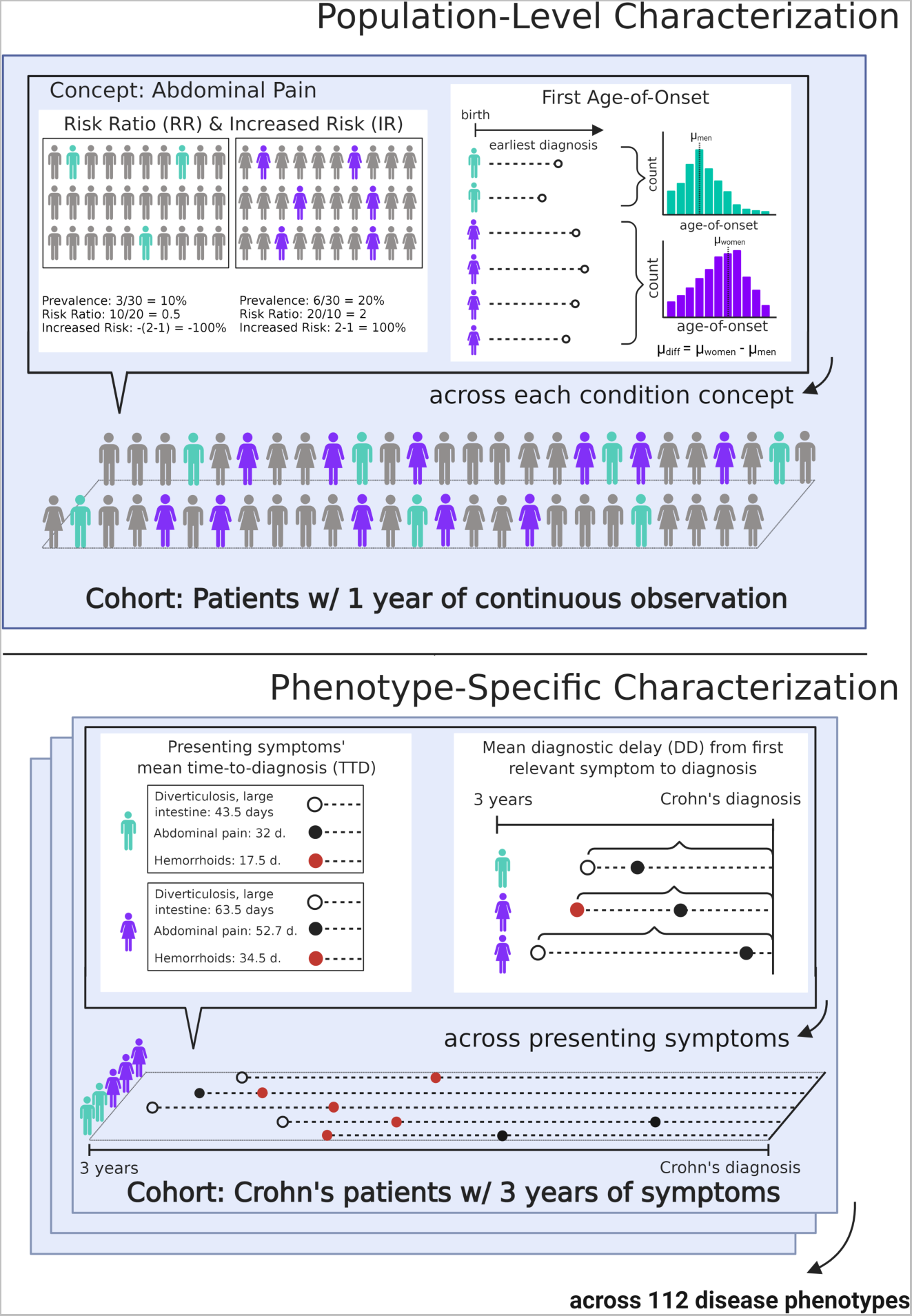
Graphical representation of methods used in the population-level characterization (top) and the phenotype-specific characterization (bottom). In the population-level characterization, we analyze diseases and conditions that occur in patients with at least one year of continuous observation. We calculate condition prevalence in women and men and compute risk ratios. We also calculate age of onset for each condition among all women and men, then estimate the mean difference in age of onset across genders. In the phenotype-specific characterization, across 112 phenotype definitions (using Crohn’s disease as an example), we aggregate the presenting symptoms prior to diagnosis in 1-, 3-, and 10-year increments for acute, mid-length, and long-term chronic conditions, respectively. We assess time to diagnosis for each presenting symptom across genders, computing the mean differences of the distributions. We also assess the diagnostic delay from first relevant symptom to diagnosis.

### Datasets

As part of the Observational Health Data Sciences and Informatics (OHDSI) initiative, clinical concepts in each de-identified dataset are standardized using the open science Observational Medical Outcomes Partnership (OMOP) common data model, which facilitates synthesis of disparate observational databases by aligning healthcare data to a uniform set of vocabularies (*15*). All condition codes are mapped to the standard vocabulary, SNOMED-CT. A breakdown of gender and demographic information for each source is shown in **Table 1**. The datasets are as follows:

**Table 1.** Summary statistics and demographic information for the four databases used in the study. Figures in the main paper are based on data from the Commercial Claims and Encounters (CCAE) database, as CCAE is the largest database by an order of magnitude – additional graphs for MDCD, MDCR, and CUIMC can be found in Supplemental Materials. Commercial Claims and Encounters (CCAE) and Medicare (MDCR) databases do not have race or ethnicity demographic information. Data from all sources span January 01, 2010 through January 01, 2020.

|  | **Claims** |  |  | **EHR** |
| --- | --- | --- | --- | --- |
| Database | Commercial Claims and Encounters (CCAE) | Medicaid (MDCD) | Medicare (MDCR) | Columbia University Irving Medical Center (CUIMC) |
| *No. of persons* | 158,388,845 | 32,806,887 | 10,252,269 | 6,747,059 |
| *No. of women* | 80,947,643 | 18,436,044 | 5,662,878 | 3,758,402 |
| *No. of men* | 77,441,202 | 14,370,843 | 4,589,391 | 2,971,591 |
| *No. of conditions* | 16,454 | 16,269 | 15,850 | 17,587 |
| *Age range (years)* | 1 - 65 | 1 - 65 | 65+ | All ages |
| *Black* | N/A | 8,897,511 | N/A | 380,926 |
| *White* | N/A | 15,828,667 | N/A | 1,069,661 |
| *Hispanic, Latino* | N/A | 2,396,864 | N/A | 529,371 |
| *Non-Hispanic, Latino* | N/A | N/A | N/A | 992,193 |

#### CCAE

The MarketScan Commercial Claims and Encounters (CCAE) dataset is curated by Truven Health Analytics, a division of IBM Watson Health (*16*). The dataset contains fully de-identified patient-level information for families covered by employer-sponsored commercial insurance (*17*). The data is derived from inpatient and outpatient medical claims as well as procedural and outpatient pharmaceutical claims. In total, the full dataset includes records from 80,947,643 women and 77,441,202 men with 16,454 unique SNOMED-CT condition codes.

#### MDCD

Medicaid in the United States is a federal program administered by states that helps reduce healthcare costs for individuals with limited income under the age of 65 (*18*). The MarketScan Medicaid Multi-State (MDCD) dataset includes healthcare claims data for individuals covered by Medicaid in 11 states, with additional demographic data such as race and disability status (*17*). It includes 18,436,044 women and 14,370,843 men with 16,269 unique SNOMED-CT condition codes.

#### MDCR

Medicare in the United States is a federal program that covers healthcare costs primarily for individuals over the age of 65 (*19*). The MarketScan Medicare Supplemental (MDCR) dataset covers retirees with employer-sponsored Medicare supplemental insurance, including claims from both the Medicare-paid portion as well as the employer-covered portion (*17*). The dataset includes information on 5,662,878 women and 4,589,391 men with 15,850 unique SNOMED-CT condition codes.

#### CUIMC

Columbia University Irving Medical Center (CUIMC) is an academic medical center that houses New York-Presbyterian Hospital and Morgan Stanley Children’s Hospital in New York City. The EHR contains inpatient and outpatient data. The CUIMC dataset includes information on 3,758,402 women and 2,971,591 men with 17,587 unique SNOMED-CT condition codes.

### Population-wide characterization

In the population-wide characterization, we aim to systematically capture patterns of condition diagnosis among women and men. Each dataset is restricted to patients with at least one year of continuous observation who had recorded encounters between January 1, 2010 and January 1, 2020. The presentation of a condition for a patient is defined by the presence of a condition code in their longitudinal health record. The earliest occurrence of a code in a patient’s longitudinal record defines the patient’s onset of diagnosis, with the date of the earliest code determining the patient’s age of onset.

All condition codes are included in the analysis, with the exception of heavily sex-specific conditions (e.g., conditions related to reproductive organs, childbirth, and pregnancy; see Methods and **Table S1** in Supplementary Materials).

The following metrics are computed for each condition in a dataset: (1) **prevalence** in women and in men (e.g., 20% of women and 10% of men have a diagnosis of abdominal pain in their record); (2) **risk ratio** based on prevalence (e.g., 0.5 for men and 2 for women for abdominal pain); (3) **increased risk** (up to -100% for men and +100% for women) to indicate whether women or men are more likely to be diagnosed with the condition; and (4) **difference in age of onset** to indicate the difference in mean age of onset between women and men (see Population-wide Characterization in **Fig. 1**). For additional information about the metrics, we provide additional information in the Methods section of the Supplementary Materials.

### Phenotype-specific characterization

In the phenotype-specific characterization, we explore how long women and men wait between presentation with relevant symptoms and eventual disease diagnosis. We describe (1) how we assess the onset of **diagnosis outcomes of interest** using clinical phenotypes, (2) how we quantify **relevant presenting symptoms** for these diagnoses, and (3) how we compute the **time to diagnosis and diagnostic delay metrics**.

#### Diagnosis Outcomes of Interest

While the population-level characterization enables an analysis of all aggregated condition codes at scale, condition codes are a noisy proxy for actual diagnosis. For the phenotype-specific analyses, disease diagnosis is measured as whether the patient’s longitudinal record satisfies a particular disease-specific set of condition criteria (i.e., a phenotype). For example, our Crohn’s disease phenotype definition requires a patient record to include any mention of the Crohn’s condition concept (SNOMED concept ID 201606) and descendants in the SNOMED-CT ontology, or alternatively mentions of concepts related to regional enteritis of the jejunum, extraintestinal Crohn’s, or complications due to Crohn’s disease (as well as these conditions’ various descendants in the SNOMED-CT hierarchy).

Phenotypes are sourced from the publicly available OHDSI Phenotype Library, which has been employed in multiple network studies using EHR and claims data (*20*, *21*). In aggregate, the 112 disease phenotypes cover a broad range of diagnosis outcomes across each ICD-10-CM disease chapter except chapters related to pregnancy, childbirth, perinatal conditions, and congenital malformations because of their sex-specific nature (**Table S4**). The phenotypes are further split according to the relative acuity of the underlying diagnosis, with different requirements for the number of years of continuous observation required prior to diagnosis corresponding to disease acuity (**Table S4**).

Given these phenotype definitions outline the diagnostic criteria for each outcome of interest, we define a patient’s diagnosis onset as their time of entry into any phenotype cohort based on their longitudinal record.

#### Relevant Presenting Symptoms

For the purposes of this paper, we consider the set of all coded condition occurrences (i.e. investigative clinical findings and other disease diagnoses) that occur at least once in a patient’s longitudinal record prior to their cohort entry as potential symptoms for a diagnosis.

In order to assess when patients first present with relevant symptoms for a given phenotype, we first asked clinical experts to manually curate a list of relevant condition codes for a subset of disease phenotypes, which we refer to as the **clinically-adjudicated relevant symptoms** (see Supplemental Methods for adjudication details). Because of the large number of disease phenotypes assessed, we separately developed a scalable approach for automatically generating relevant symptom lists, which we refer to as the **algorithmically-generated relevant symptoms** (see Supplemental Methods for generation details). We validated our algorithmically-generated symptom lists against the gold-standard set of clinically-adjudicated symptoms and found that the algorithmically-generated symptoms are able to consistently and accurately recall the majority of the clinically-adjudicated symptoms, supporting our usage of the algorithmically-generated relevant symptoms across our analyses (see Validation of Algorithmically-Generated Symptoms in Supplemental Methods and **Table S2** for precision/recall/F1 results).

In all cases, we define the time of onset for a given symptom as the earliest date the symptom is recorded within the continuous observation period in a patient’s longitudinal record prior to diagnosis.

#### Metrics for time to diagnosis and diagnostic delay

To capture different temporal aspects of diagnosis, we calculate and report (1) **symptom-based time to diagnosis** (abbreviated TTD) and (2) **patient-centric diagnostic delay** (abbreviated as DD).

The **symptom-based time to diagnosis** metric measures how much longer women have to wait compared to men that present with the same relevant symptom before receiving the same disease diagnosis (see left-hand side of the Phenotype-Specific Characterization diagram in **Fig. 1**). In other words, across relevant symptoms in a disease phenotype, we calculate and report the mean time to diagnosis differences for women and men presenting with sets of relevant symptoms.

The **patient-centric diagnostic delay** metric measures how long individual patients have to wait from their first relevant symptom until disease diagnosis (see righthand side of the Phenotype-Specific Characterization diagram in **Fig. 1**). The diagnostic delay is symptom-agnostic and represents when patients could be suspected of having a particular disease diagnosis. To assess statistically significant differences in diagnostic delay between women and men, we apply the Kolmogorov-Smirnov (KS) to test our null hypothesis that there is no difference in the cumulative distributions of diagnostic delay for women and men (see Methods in Supplementary Materials for more details) (*23*).

## Results

Descriptive statistics for each dataset are shown in **Table 1**, with the total dataset representing 208,177,994 patients with longitudinal records. The companion website for this study (https://even.dbmi.columbia.edu/characterization) provides interactive visualizations of the results described here to allow researchers to further explore differences in the patterns of disease diagnosis between women and men.

### Population-Wide Characterization

#### Risk Ratio

Among the two largest claims databases (CCAE and MDCD), the risk ratio distributions are asymmetrically distributed and indicate that women are more at risk of receiving a diagnosis across conditions (**Fig. 2**, left-most column: in CCAE and MDCD respectively, there are 7,327 and 6,992 conditions with risk ratio >1 for women compared to 5,972 and 5,509 conditions with risk ratio >1 for men). When considering the tails of these risk ratio distributions, for conditions where women or men are significantly more at risk (i.e. where women’s risk ratio is greater than 3 or less than 1/3), we find that there are more conditions where women in CCAE and MDCD experience a greater risk (**Fig. 2**, left-most column: in CCAE there are 643 conditions where women are more than 3 times at risk than men, compared to 416 conditions where men are more than 3 times at risk; similarly, in MDCD, this is 548 conditions for women vs 309 conditions for men). In the MDCR claims dataset, the risk ratio distributions show approximately equivalent risk distributions across conditions between women and men, while the CUIMC EHR dataset conversely shows that men are more at risk for most conditions (e.g. there are 430 conditions where men are more than 3 times at risk, while only 234 conditions where women are more than 3 times at risk).

**Fig. 2.**
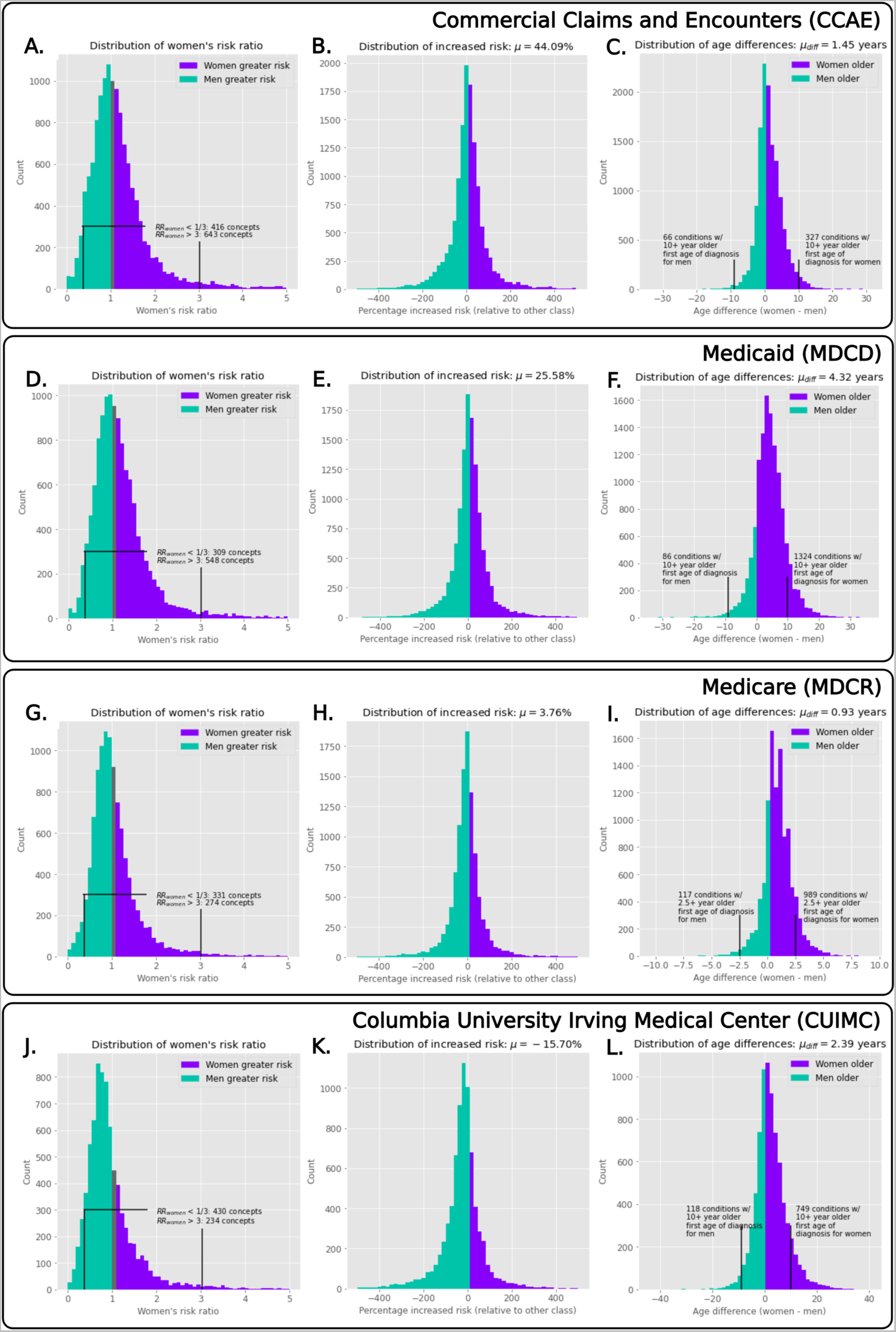
Population-level distributions showing women’s risk ratios, increased risk, and age of onset differences. We visualize the women’s risk ratio distributions for the various datasets (left-most column), where we see that, in the largest two databases, women are more likely to be more at risk (as shown in the differences in counts). Considering the increased risk (middle column) for all datasets except CUIMC, women were at increased risk for all conditions. When we examine age of onset distributions (right-most column), women are consistently older when first diagnosed for all conditions across all datasets.

#### Increased Risk

Distributions of increased risk for all conditions are illustrated by data source in Fig. 2 (middle column). Increased risk is consistently positive across all claims databases (CCAE, MDCD, MDCR), indicating that women are systematically more likely to be diagnosed with most conditions across these databases (range: 3.76% to 44.09%). Consistent with the previous risk ratio findings, the EHR dataset (CUIMC) displays an opposite trend where men on average are 15.70% more likely to be diagnosed with most conditions.

#### Age of Onset

When examining distributions of differences in age of onset (**Fig. 2**, right-most column), women are older than men when first diagnosed across all conditions and datasets (0.93 to 4.32 years older on average at onset). Overall age differences are smaller in MDCR because of age-censoring, as the dataset only includes patients that are at least 65 years of age who qualify for Medicare. When we additionally consider conditions where the mean difference in age of onset is greater than 10 years, we find a larger number of conditions exist where women are over a decade older at diagnosis onset compared to the number of conditions in men (e.g. in CCAE there are 327 conditions where women are a decade older than men versus 66 conditions where men are a decade older than women, in MDCD it is 1,324 for women and 86 for men, and for CUIMC it is 749 for women and 118 for men). Given the older age of patients in the MDCR dataset, we examined conditions where the mean difference in age of onset was greater than 2.5 years, and find that in MDCR there were 989 conditions for which women were 2.5 years older versus 117 for which men were 2.5 years older.

### Phenotype-Specific Characterization

Across presenting symptoms, acute and chronic phenotypes, and databases, we find that women systematically experience longer time-to-diagnoses and diagnostic delays compared to men. For the purposes of narrative clarity, figures in the main-text display findings based on the largest claims (CCAE) dataset, with additional figures from the other three datasets (MDCD, MDCR, and CUIMC) shown in the Supplementary Materials (**Figs. S1-6** and **Table S5**).

#### Time to Diagnosis (TTD) Differences

Across datasets, women consistently experience longer time to diagnoses (TTDs) than men. After matching patients based on the set of algorithmically-generated relevant symptoms, women with any given relevant symptom are more likely to experience a longer time to diagnosis than men with the same symptom (e.g. in CCAE, 86.3% of the symptoms for the acute phenotypes led to a later diagnosis for women, 83.2% for the mid-length chronic phenotypes, and 69.6% for the long-term chronic phenotypes; see **Table S5** for breakdowns across datasets). When we quantify the magnitude of women’s time-to-diagnosis differences across relevant symptoms, women consistently experience longer TTDs across CCAE, MDCD, and MDCR. Across the relevant symptoms, women in CCAE must on average wait 8.0 days longer than men across acute phenotypes, 25.3 days for the mid-length chronic phenotypes, and 51.9 days for the long-term chronic phenotypes (see **Table S5** for breakdowns for each database). In **Fig. 3** we visually illustrate mean time to diagnosis differences for each individual phenotype assessed in CCAE and note that women consistently experience longer TTDs in 110 of the total 112 phenotypes analyzed. Across the other large claims databases, we note similar trends in TTD differences (MDCD in **Fig. S1**: 104 of the 112 phenotypes, MDCR in **Fig. S2**: 107 of the 112 phenotypes), although the trend is less readily apparent for the EHR dataset (CUIMC in **Fig. S3**).

**Fig. 3.**
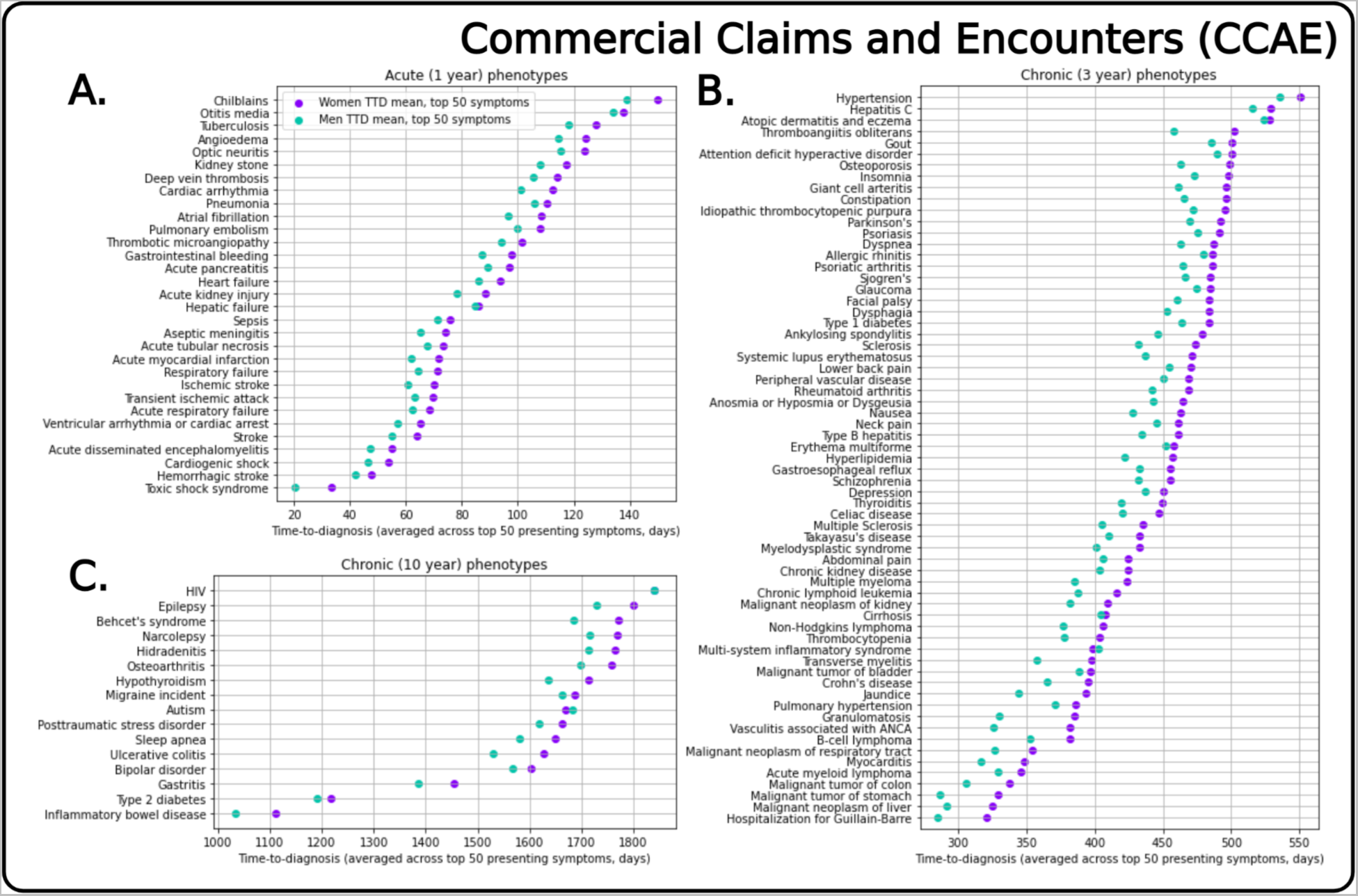
Mean time to diagnosis (TTD) for women and men across all phenotypes in the Commercial Claims and Encounters (CCAE) dataset. Aggregated results show mean TTDs broken down by phenotype for CCAE, the largest private-insurance claims database. Each X-axis shows the mean TTD difference averaged across TTDs for the top 50 presenting symptoms . Visualization of TTD illustrates that women are consistently diagnosed later than men when with a common set of presenting symptoms, when we consider the same relevant conditions for all phenotypes (except in the case of autism, HIV and multi-system inflammatory disorder). This consistent trend persists across MDCD and MDCR (Figs. S1, S2), indicating that these findings persist regardless of whether a patient had private or government insurance.

When we assess the mean time-to-diagnosis using clinician-adjudicated relevant symptoms, we observe results consistent with the algorithmically-generated relevant symptoms. In both cases, across phenotypes assessed, women experience longer time-to-diagnoses than men (**Table S3**). In fact, in all but one phenotype, using the clinician-adjudicated relevant symptoms revealed even larger differences in time to diagnosis. For additional information about comparisons between the clinician-adjudicated relevant symptoms and the algorithmically-generated symptoms, see Validation of Algorithmically-Generated Symptoms in the Supplemental Methods.

To allow researchers to explore and expand on our findings, we also publicly present interactive visualizations that depict the individual distributions of relevant symptoms for each phenotype and each dataset (**Fig. 4**). These online, accessible visualizations notably make it easy to see that, across relevant symptoms, phenotypes, and datasets, the time between initial presentation and receipt of diagnosis is generally longer for women than for men.

**Fig. 4.**
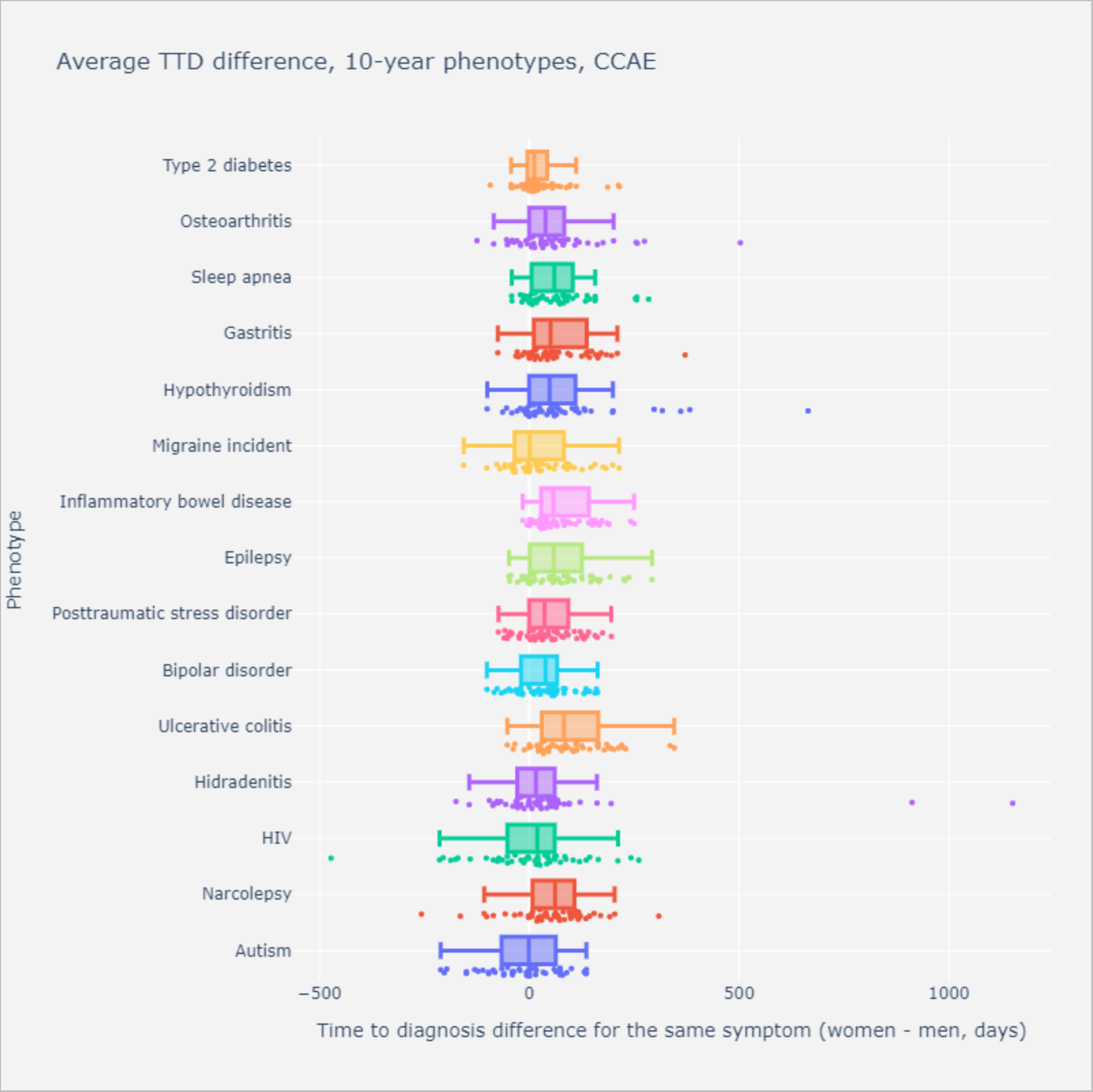
Mean time to diagnosis (TTD) differences using the top-50 presenting symptoms for women and men in a subset of 10-year chronic phenotypes from the Commercial Claims and Encounters (CCAE) dataset. Each dot represents the TTD difference between women and men for the same matched symptom; a positive difference indicates that women have to wait longer than men. Note how it takes longer until diagnosis for the vast majority of women’s presenting symptoms. Also, when we consider mean TTD based on the top-50 presenting symptoms (shown as the bar graphs), the overall TTD is consistently longer for women than for men. These trends persist across databases and phenotypes.

#### Diagnostic Delay (DD) Differences

Across the claims datasets (CCAE, MDCD, MDCR), there are significant gendered differences in diagnostic delay for almost all phenotypes. Considering CCAE, there are significant gendered differences in diagnostic delay (DD) for the acute (31 of 31), mid-length chronic (62 of 65), and long-term chronic (15 of 16) phenotypes with significance threshold of p < 0.01 (**Fig. 5**). Women on average must wait 21.0, 62.9, and 134.0 days longer than men after their first presentation with any relevant symptom for the acute, mid-length chronic, and long-term chronic phenotypes respectively (**Table S5**). These trend persists across MDCD and MDCR data regardless of phenotype course (i.e., acute vs. chronic), with most phenotypes having statistically significant differences with longer observed diagnostic delays for women (**Figs. S4-S5**). The diagnostic delay differences are less readily apparent for CUIMC data (i.e., fewer phenotypes were statistically significant, **Fig. S6**).

**Fig. 5.**
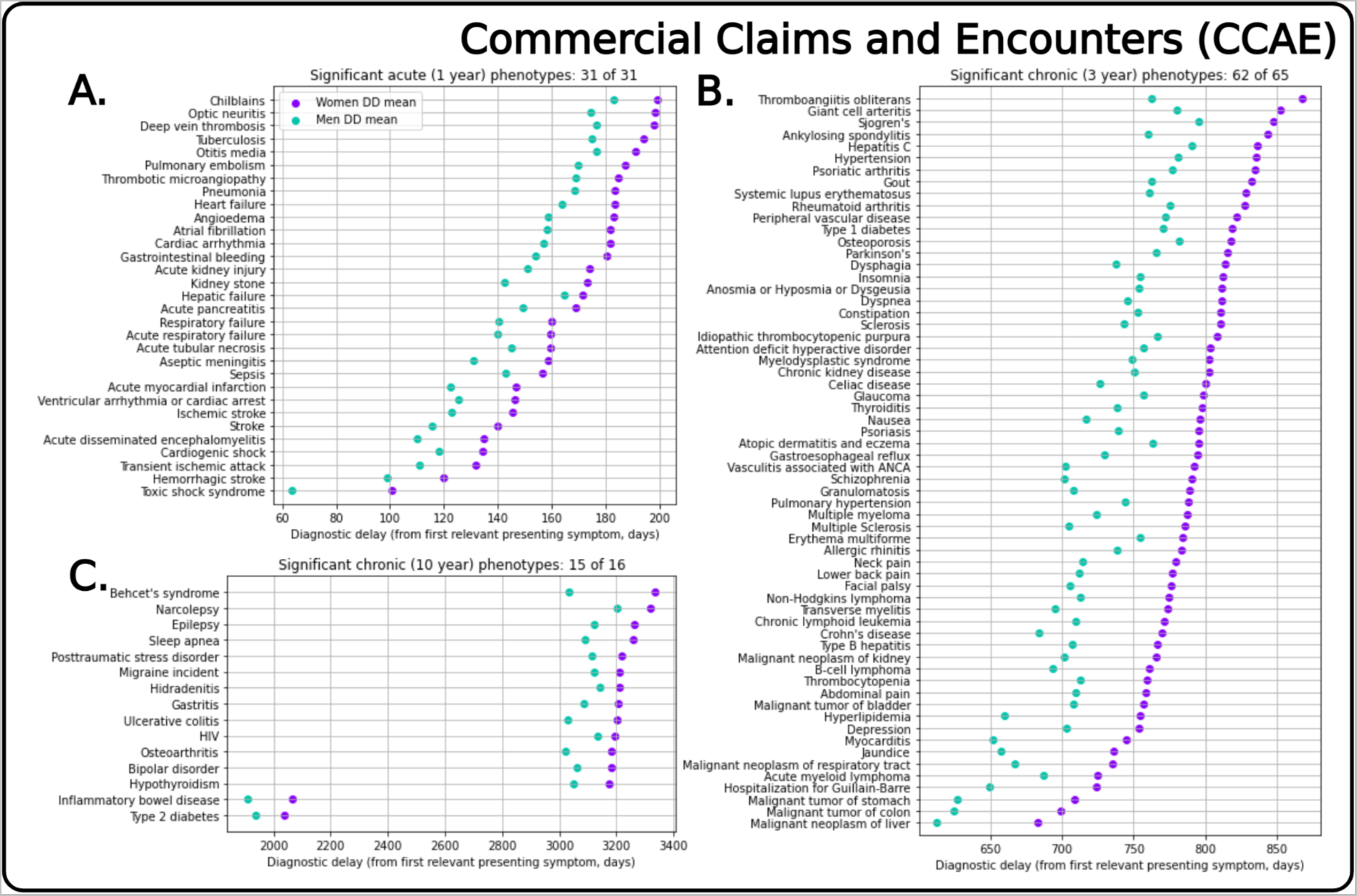
Mean diagnostic delay (DD) for women and men across phenotypes in the Commercial Claims and Encounters (CCAE) dataset. Aggregated results show DD for women and men after performing Kolmogorov-Smirnov tests to calculate if the difference in distributions for women and men was statistically significant (based on threshold of p < 0.01). Visualizing the DD (i.e., the time from first relevant symptom to time of diagnosis) illustrates that, for all significant differences, women have to wait longer from first relevant symptom presentation to diagnosis.

## Discussion

By leveraging the OHDSI federated research platform (*15*) and the OMOP common data model (*32*), this study represents one of the first systematic, comprehensive, large-scale characterizations of disease diagnosis differences across genders. Where previous studies rely on highly curated, single-disease cohorts which include up to a few thousand patients (*24–30*), our systematic characterization considers over 208 million patients across more than a hundred acute and chronic diseases. This study’s heterogenous study population includes geographically diverse patients covered by both government and private insurance, with longitudinal records spanning more than a decade from both claims and EHR datasets. To date, the only comparable large-scale characterization of gender differences in diagnosis is limited to a single longitudinal Danish national health registry study that examines age of onset and prevalence differences (*31*). Taken together, the findings from this study indicate that systematic differences exist in disease diagnosis across genders, with these differences persisting across conditions and datasets.

While we expect sex-specific conditions to be diagnosed differently across genders, our findings suggest that women and men are differently diagnosed for a much broader set of conditions. In the population-wide characterization, across all datasets women were observed to be at greater risk of receiving diagnoses for most conditions in the general population. Despite being more at risk, women also were older than men when first diagnosed, aligning with previous findings from the longitudinal Danish study (*31*). In the phenotype-specific characterization, we additionally observe staggering differences in time-to-diagnosis and diagnostic delay between women and men. After examining relevant symptoms leading up to diagnosis, our findings indicate that it takes longer for women to be formally diagnosed, even when comparing across a common set of relevant symptoms. Likewise, across datasets, women experience significantly longer diagnostic delays compared to men. These findings suggest that differences in the timeliness of diagnosis are due to more than differential disease presentation, as the analyses use similar sets of relevant symptoms. Indeed, given that diagnostic criteria are traditionally evaluated based on men’s symptom etiology, our analysis is likely a conservative estimate of time to diagnosis and diagnostic delay differences (since in cases where women and men differ in disease presentation, women may be even less likely to have received access to a diagnosis).

Overall, the findings have large implications regarding gender bias in medicine, highlighting how neither previously assumed equality nor assumed differences between women and men are necessarily appropriate (*33*). With women being more at-risk and older when presenting with conditions in a population (population-level characterization), but also experiencing longer time to diagnoses and diagnostic delays (phenotype-level characterization), our findings raise important questions concerning how care delivery is impacted by both genuine biological sex differences and potential gender biases in care. The consistent nature of these time to diagnosis and diagnostic delay differences suggest that critical differences in initial presentation may not be fully represented in the claims data. In light of these findings, the potential for encoding algorithmic bias in models trained on clinical data also merits special attention, especially as artificial intelligence proliferates in medicine. As women are consistently being diagnosed later than men, even when they present with similar symptoms, there is a genuine risk of erroneously and automatically down-weighting the symptoms of women during model training. Further, historical patterns of delay and lack of attention to earlier symptom presentation in women may also result in learned assumptions of longer lag times in disease progression that further exacerbate differences by prioritizing earlier treatment in men. These results suggest that the individual information in presenting symptoms may be different, and warrants a deeper dive into what drives these observed differences. We emphasize that our approach can be easily extended to new disease phenotypes or reproduced across datasets, since we make all OHDSI phenotypes and analytic code available to the public to foster additional characterizations at scale.

Our findings should be considered in light of several potential limitations. First, we recognize the importance of how other demographic factors could be health determinants, and note the potential insights that could be additionally afforded by applying an intersectional lens to these analyses (e.g., examination along subdivisions of both gender and race). Unfortunately, limitations in the data sources did not allow for this type of analysis. Nonetheless, we posit that comparison of diagnostic patterns between and among subgroups would find similarly notable or perhaps even more extreme differences given prevailing literature on inequities in care delivery. Second, while systematic and comprehensive, our approach is care setting agnostic and does not distinguish between visits in the outpatient or inpatient setting, nor does it stratify by provider specialty. Nonetheless, we note that the differences observed are both consistent and staggering, and we posit that since these trends exist in aggregate data, they would likely persist irrespective of care setting. Third, in our phenotype analyses, we used algorithmically-generated relevant symptoms to scale our analyses across the numerous phenotypes. As we noted in the results, after extensive validation, using clinician-adjudicated relevant symptoms produced even more staggering results than using the algorithmically-generated relevant symptoms. Clinical expertise essentially filtered out anomalous symptoms and focused on highly predictive symptoms. Since the majority of presenting symptoms had longer time to diagnoses for women, clinical filtering focusing on highly predictive symptoms made the findings even more poignant. Finally, we sought to minimize potential biases attributed to variability in healthcare utilization and access. We required all patients to meet the same minimum number of years of continuous observation and performed our analyses independently on each data source without aggregation, to preserve differences in insurance status (private or government plan) and data provenance (claims or EHR). Our analysis found trends in claims data that were consistent regardless of insurance type. Many of those same trends were also evident in the EHR data, but to a varying degree.

In conclusion, while we cannot yet causally explain why these differences occur, the presence of these consistent, staggering differences in diagnosis patterns suggest that women and men in both public and private insurance are differentially diagnosed (and potentially differentially treated) after presentation with symptoms. The findings from this paper are the first step toward systematically identifying sex and gender-disparities in disease diagnosis, and suggest that we must further examine these differences and identify on a per-disease basis how and why these systematic differences occur.

## Acknowledgments

We would like to thank the members of the Elhadad lab at the Columbia Department of Biomedical Informatics, the clinical experts at Johnson & Johnson who reviewed and validated the presenting symptoms, and BioRenderer for graphics.

## Author contributions

Conceptualization: TYS, PBR, NE

Methodology: TYS, PBR, NE

Investigation: TYS, JH, HRN

Visualization: TYS

Funding acquisition: PBR, RC, NE

Project administration: NE

Supervision: NE

Writing – original draft: TYS, HRN, NE

Writing – review & editing: TYS, HRN, RC, PBR, NE

## Data and materials availability

Data used in this study are from standardized OMOP version of electronic health records and claims datasets. While the data are not publicly available, all raw summary results of the analysis are available upon request. Interactive visualizations of all study results are available on the companion website of the study at https://even.dbmi.columbia.edu/characterization. Code for analysis is available at https://github.com/elhadadlab/gender-differences-characterization.

## Footnotes

1 As defined by the World Health Organization, gender is as a social construct while sex is a biological variable (1). We use gendered terms (women, men) throughout this study. Furthermore, because we carry out analyses on large healthcare databases that encode gender as a binary variable only, our analysis is limited to this binary definition.

