## Supplemental Figures for "Large-scale characterization of gender differences in diagnosis prevalence and time to diagnosis"

### Materials

*Patient Gender*

The OMOP common data model uses a binary representation of gender (Male, Female), and thus only these categories were considered in the analyses. Gender categories from CUIMC were extracted from the electronic health record's gender field, with the fields originally collected from patient self-reporting. For the CCAE, MDCD, and MDCR claims datasets, gender was collected from patient self-reporting when patients were initially enrolled in the datasets.

*Conditions Included in Population-level characterization*

We excluded rare conditions and sex-specific conditions in our characterization analyses. Rare conditions were operationalized as any condition with less than 50 total occurrences in a given database's population. Sex-specific conditions were identified through three complementary approaches: (1) by leveraging OHDSI's DataQualityDashboard set of gender-specific concepts (*26*), (2) by identifying concepts that were more than 99% prevalent among a single gender in database, and (3) by manually filtering out condition names that contain references to sex-specific anatomical terms (see **Table S3** for full list of anatomical terms).

*Phenotypes Included in Phenotype-specific characterization*

The 112 disease phenotypes considered in the phenotype-specific characterization are listed in **Table S1**. These disease phenotypes cover all ICD-10-CM chapters besides chapter 15 (pregnancy, childbirth and the puerperium), chapter 16 (certain conditions originating in the perinatal period) and chapter 17 (congenital malformations, deformations and chromosomal abnormalities) because of their sex-specific nature.

The phenotypes also cover different disease acuity levels. Phenotypes were split into acute diseases (31 phenotypes), mid-term chronic diseases (65 phenotypes), and long-term chronic diseases (16 phenotypes). For the acute diseases, time-to-diagnosis metrics were computed across a 1-year lookback period, while for the mid-term chronic diseases, we used a 3-year lookback period. Finally, for the long-term chronic diseases, a 10-year lookback period was used.

All phenotype definitions are available in OHDSI-compliant JSONs structure on GitHub and are reproducible on any OMOP-compliant observational health database (*27*).

*Study Approval*

The study protocol was reviewed and approved by the Columbia University Irving Medical Center Institutional Review Board.

### Methods

*Population-level characterization: Increased Risk Calculations*

Because risk ratios are centered around 1 and bounded between 0 and positive infinity, a risk ratio distribution is naturally skewed and more sensitive toward large, positive risk ratio values. To address this, we instead use a zero-centered, symmetrical "increased risk" metric that we based on the original risk ratio.

To calculate the increased risk for a particular condition, determine whether the women (or men's) risk ratio is greater for that given condition. For instance, if a women's risk ratio of 1.25 for a given condition indicates that women are 1.25 times at risk to obtain the diagnosis code than men, the men's risk ratio for the same condition would be 0.8. A women's risk ratio for any condition is the inverse of the men's risk ratio. Then, subtract 1 from the larger of the risk ratios (in this case, 1.25 – 1, leaving us with the proportion of increased risk), and multiply by negative one if the risk ratio for men was the greater value. The resultant value is thus centered uniformly around 0, with positive or negative values corresponding to women (or men) being more at risk. The magnitude of the value corresponds to how much more at-risk a particular gender would be. In formula form, the metric is defined as:

If $RR_{men} > RR_{women}$, increased risk = $-1 * (RR_{men} - 1)$,

Else if $RR_{women} > RR_{men}$, increased risk = $RR_{women} - 1$

*Phenotype-level characterization: Algorithmically-Generated Relevant Symptoms*

Because of the large number of disease phenotypes assessed, we separately developed a scalable approach for automatically generating relevant symptoms. As noted in the main-text, we consider the set of all coded condition occurrences (i.e. investigative clinical findings and other disease diagnoses) that occur at least once in a patient's longitudinal record prior to their cohort entry as potential symptoms for a diagnosis. Symptoms that only present in a single gender are filtered out, to focus the analysis on symptoms that occur in both women and men, controlling for potential sex-differences in disease presentations. To algorithmically quantify which symptoms are relevant to a given phenotype, we rank presenting symptoms according to their term frequency-inverse document frequency (tf-idf) score by treating each occurrence of a presenting symptom as a term and each separate disease phenotype as a document (*22*). The application of tf-idf enables us to separate relevant symptoms (e.g., anemia ranks high for the Crohn's disease phenotype) from symptoms that are prevalent among many phenotypes which are not predictive for diagnosis (e.g., cough is common across almost all phenotypes).

*Phenotype-level characterization: Validation of Algorithmically-Generated Symptoms*

To validate the usage of algorithmically-generated relevant symptoms, we compared their performance against clinician-adjudicated symptoms as gold-standard labels. We selected seven long-term chronic phenotypes (HIV, Crohn's disease, Hidradenitis suppurativa, Osteoarthritis, Chronic gastritis, Epilepsy, and Migraine) and had clinical experts identify among the top-100 symptoms the clinically relevant symptoms to the disease. For instance, "abdominal pain" was annotated as relevant to the diagnosis of Crohn's disease, while "depression" was annotated as not relevant to the diagnosis of epilepsy (even if it is a common co-morbidity). Clinicians were also asked if any crucial diagnosis symptoms were missed. Given these gold-standard symptom annotations, the precision/recall/F1 scores for the top-50 and top-100 symptoms were calculated and reported in **Table S2**.

For all seven phenotypes, the top-50 algorithmically-generated symptoms had a consistently higher precision than the top-100 algorithmically-generated symptoms, indicating that the ranked order of symptoms did correspond to how important symptoms were for disease diagnosis. Many symptoms flagged as false positives were noted by clinicians as common comorbidities (such as "depressive disorder" for the epilepsy phenotype) or potentially relevant symptoms (such as "hyperlipidemia" for the hidradenitis phenotype, due to associated metabolic syndrome). Given the consistently higher precision values and better F1 scores using the top-50 symptoms across all phenotypes, we decided to use the top-50 algorithmically-generated relevant symptoms throughout our analyses.

Further, as noted in the main-text, the time-to-diagnosis results using the clinician-adjudicated relevant symptoms (results reported in **Table S3**) were consistent with the findings using the algorithmically-generated relevant symptoms. In all but one case, women were even more likely to experience a longer time-to-diagnosis using the clinician-adjudicated relevant symptoms. Clinical filtering essentially removed anomalous symptoms, and since the majority of symptoms are diagnosed later in women, removing anomalous symptoms kept the overall direction of the findings consistent (e.g. women still experienced longer time-to-diagnoses in all the cases using the clinician-adjudicated relevant symptoms).

*Phenotype-level characterization: Calculating Time-To-Diagnosis Means*

For each presenting symptom, we quantify the mean and standard deviations for how long women and men have to wait between presentation with that particular presenting symptom and disease diagnosis (defined as cohort entry into the phenotype). We calculate differences between women and men by subtracting a particular symptoms' mean for the men's time-to-diagnosis from women's mean time-to-diagnosis. To calculate the aggregate measures that we plot in **Fig. 4** and **Figs. S1-S3**), we average the mean time-to-diagnoses for women and men across all top-50 algorithmically-generated relevant symptoms. In other words, the TTD graphs in **Fig. 4** and **Figs. S1-S3** averages the time-to-diagnoses across the top-50 algorithmically-generated relevant symptoms for each phenotype.

*Phenotype-level characterization: Calculating Diagnostic Delay*

For **Fig. 6** and **Figs. S4-S6** we calculate the diagnostic delays for women and men. The diagnostic delay is defined as the time between first relevant symptom (from first presentation of any among the set of top-50 algorithmically-generated relevant symptoms) and disease diagnosis (defined as cohort entry into the phenotype). This diagnostic delay metric is consistent with existing literature, where most calculations of diagnostic delay rely on patient self-reporting (e.g., for osteoarthritis diagnostic delay in Canada being from first relevant pain presentation to diagnosis (*35*)).

*Phenotype-level characterization: Computing Significance for Diagnostic Delays with Kolmogorov-Smirnov (KS) Tests*

**Fig. 6** and **Figs. S4-S6** show the diagnostic delays where the differences between women and men are statistically significant. In order to calculate significant differences, we first plot the cumulative distributions of diagnostic delays for women and for men. We then utilize SciPy to calculate the KS difference across these two cumulative distributions (*36*); the KS test is a nonparametric test for whether two samples differ from each other at a particular significance level (*29*). We use a p-value cutoff of 0.01 and plot the results of diagnostic delays that are significant at that level.

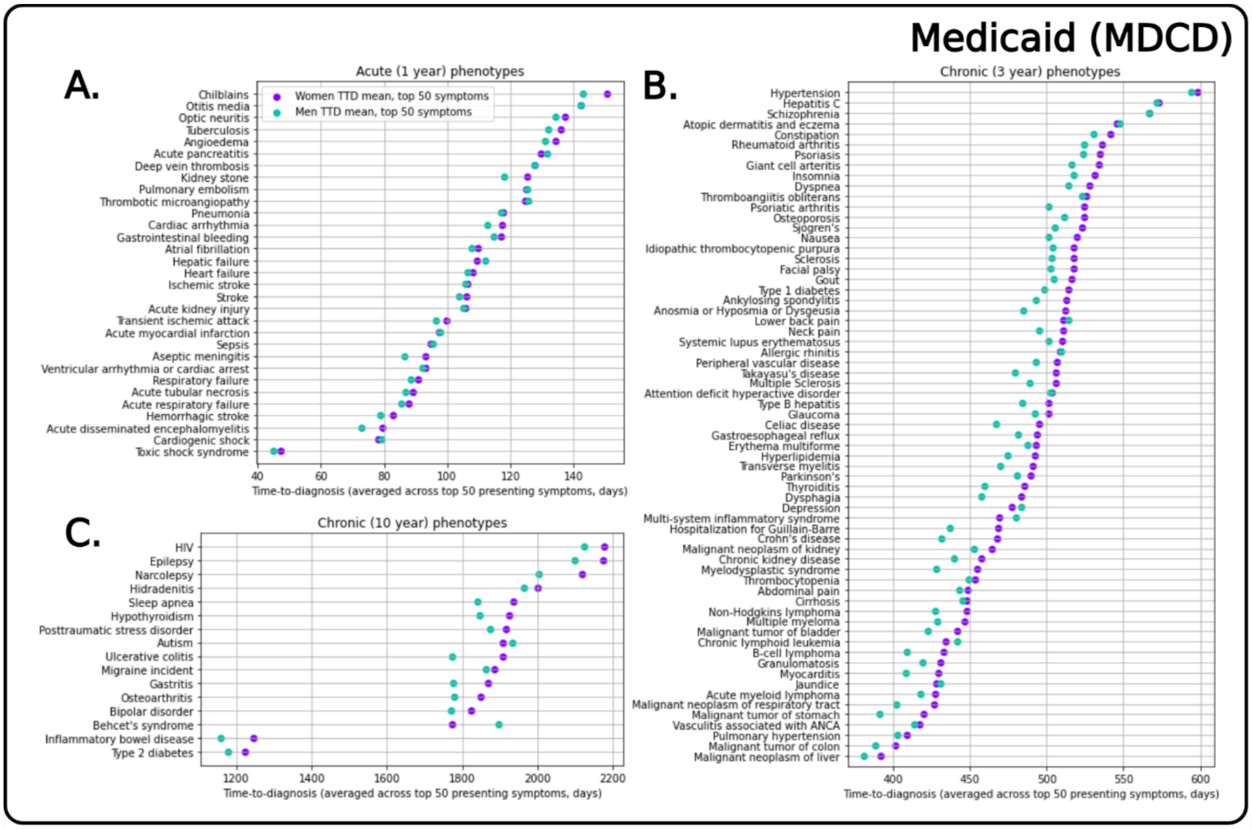


**Fig. S1. Mean time-to-diagnosis (TTD) for women and men across all phenotypes in the Medicaid (MDCD) dataset.** Aggregated results show mean TTDs broken down by phenotype for Medicaid. Visualizing TTD for women and men shows that, for most phenotypes, women are consistently diagnosed later than men when we evaluate mean TTD based on the top-50 symptoms for each phenotype. Acute pancreatitis, lower back pain, depression, multi-system inflammatory disorder, chronic lymphoid leukemia, jaundice, and Behçet's syndrome are the only phenotypes where men experience longer TTDs.

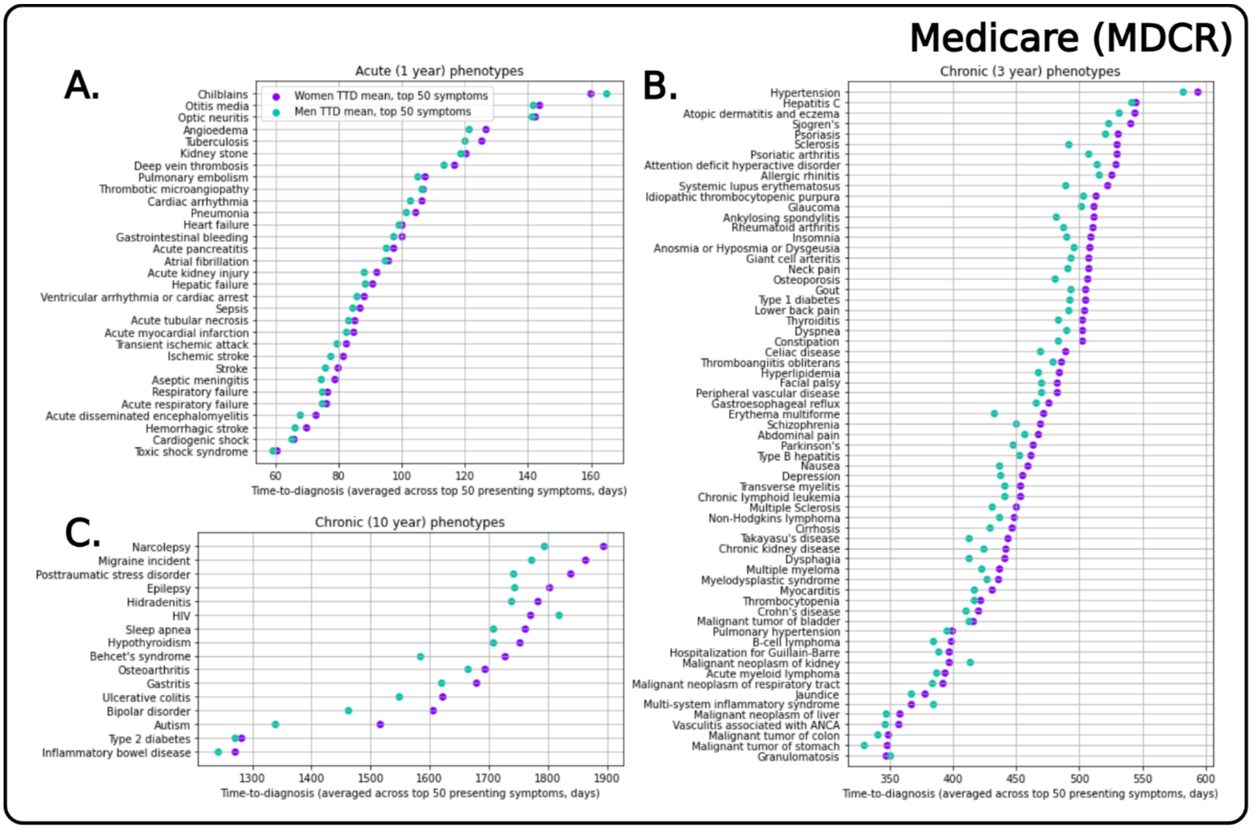


**Fig. S2. Mean time-to-diagnosis (TTD) for women and men across all phenotypes in the Medicare (MDCR) dataset.** Aggregated results show an even more marked trend in mean TTD differences broken down by phenotype. Visualizing the TTD means for MDCR clearly shows women being consistently diagnosed later than men for almost all phenotypes except chilblains, malignant neoplasm of kidney, multi-system inflammatory syndrome, granulomatosis, and HIV.

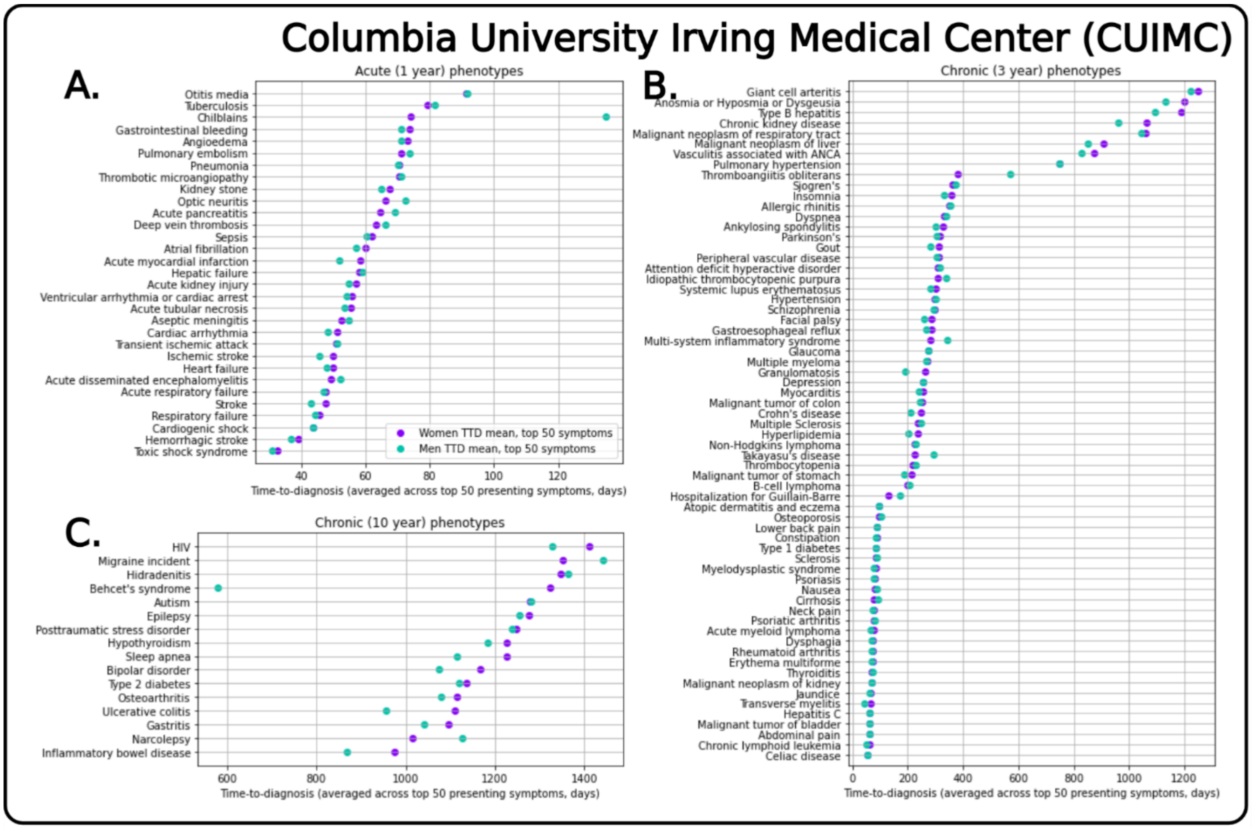


**Fig. S3. Mean time-to-diagnosis (TTD) for women and men across all phenotypes in the Columbia University Irving Medical Center (CUIMC) dataset.** Aggregated results showing the mean TTDs for women and men broken down by phenotype for CUIMC. Visualizing the TTD, we note there are a number of phenotypes where women are diagnosed later than men as well as phenotypes that show otherwise. For most phenotypes, TTD differences between women and men are small.

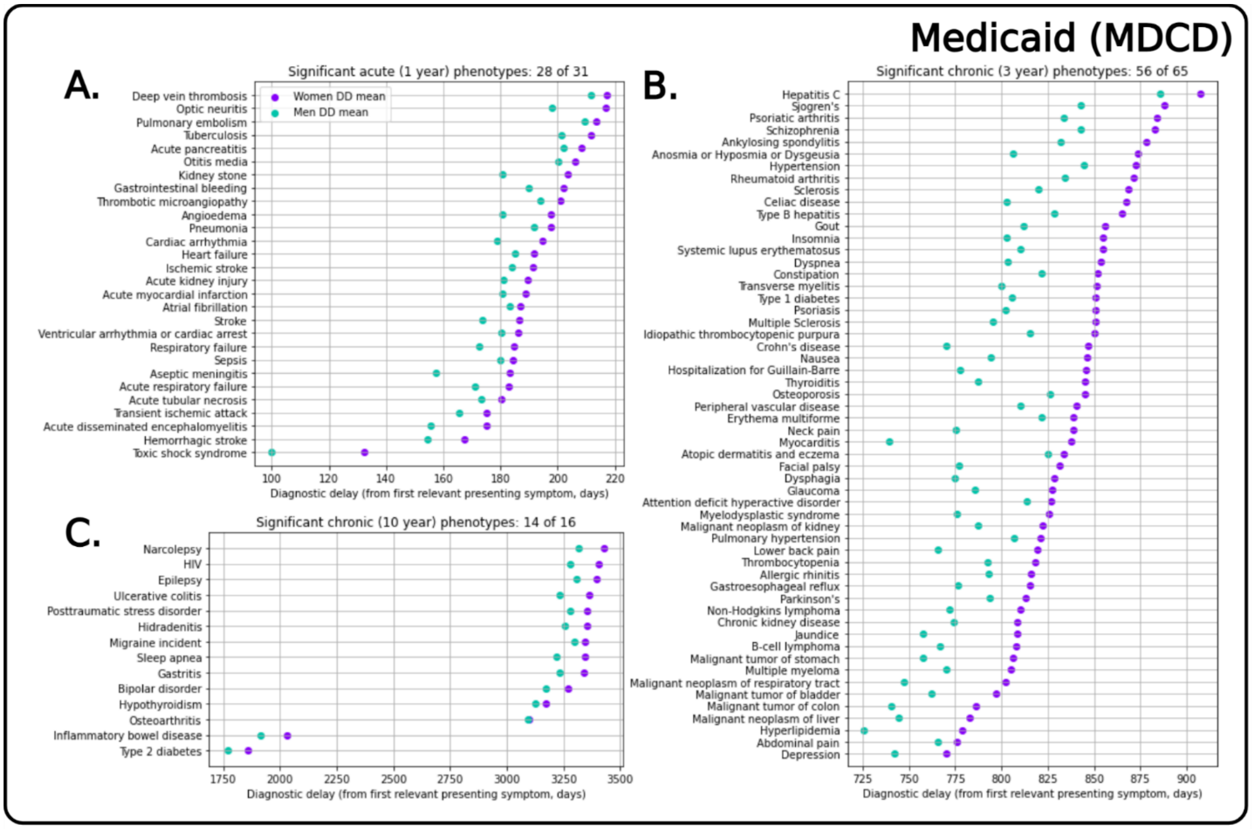


**Fig. S4. Mean diagnostic delay (DD) for women and men across phenotypes in the Medicaid (MDCD) dataset.** Aggregated results show DDs for women and men after performing the KS-test to determine if the difference in distributions was statistically significant using p < 0.01 as threshold. Visualizing the DD for women and men illustrates that, for most significant phenotypes, women consistently have to wait longer from first relevant symptom presentation to diagnosis.

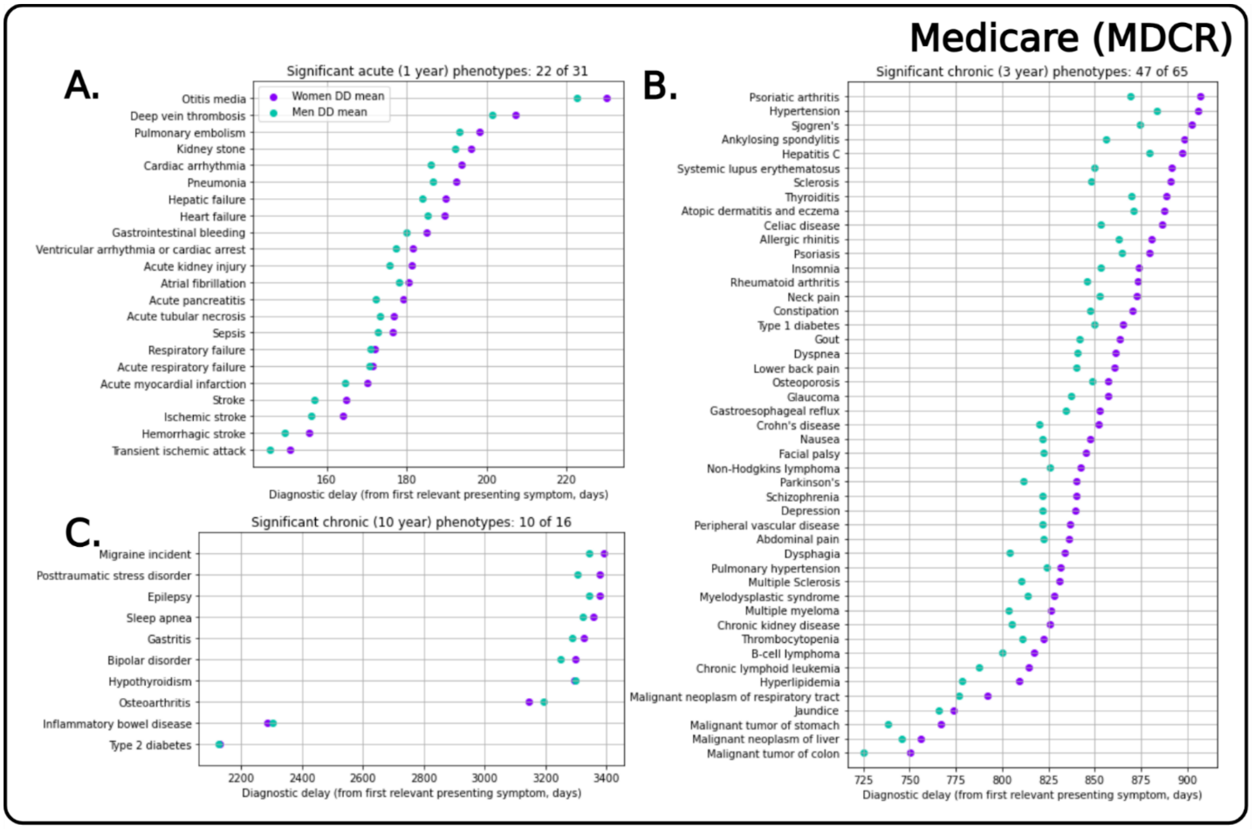


**Fig. S5. Mean diagnostic delay (DD) for women and men across phenotypes in the Medicare (MDCR) dataset.** Aggregated results show DDs for women and men after performing KS-test to determine if the difference in distributions was statistically significant using $p < 0.01$ as threshold. Visualizing the DD for women and men illustrates that, for most significant phenotypes, women consistently have to wait longer from first relevant symptom presentation to diagnosis.

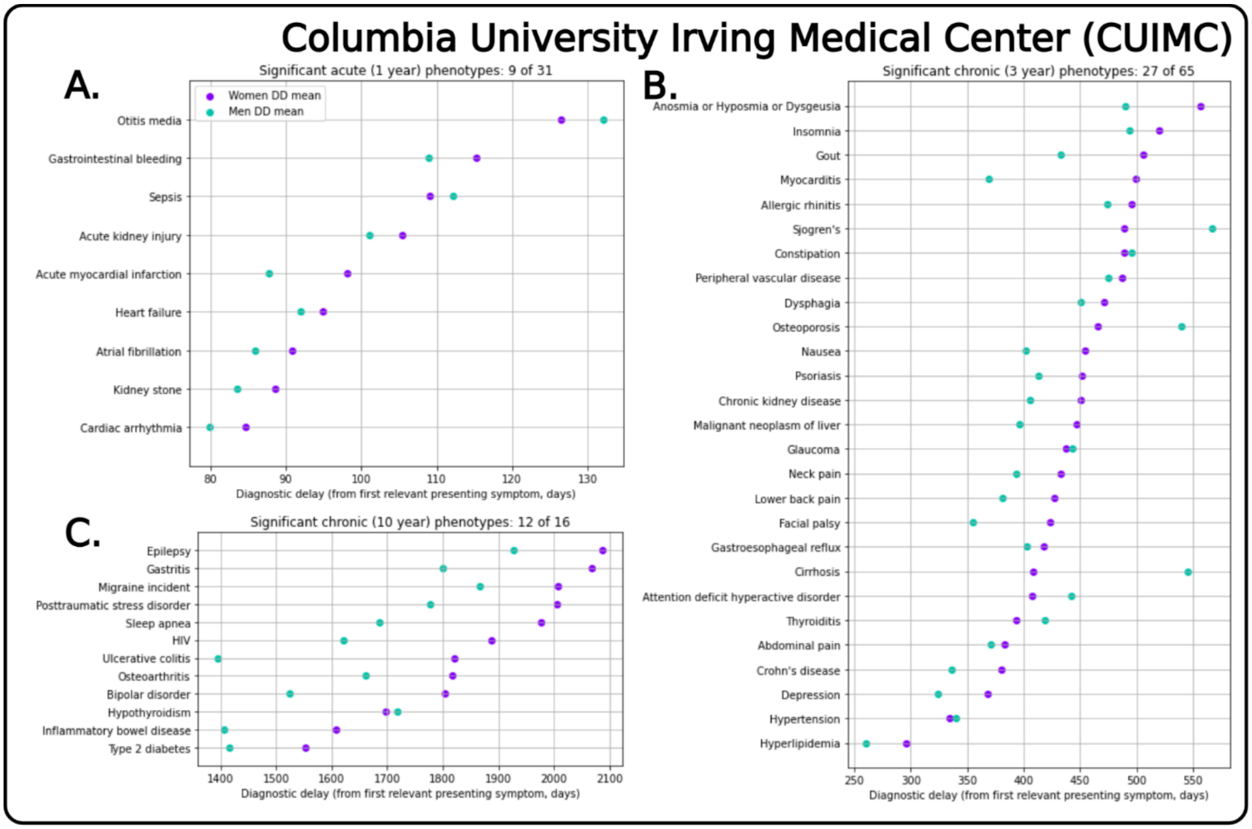


**Fig. S6. Mean diagnostic delay (DD) for women and men across phenotypes in the Columbia University Irving Medical Center (CUIMC) dataset.** Aggregated results show DDs for women and men after performing KS-test to determine if the difference in distributions was statistically significant using $p < 0.01$ as threshold. Visualizing the DD for women and men shows that there are fewer significant diagnostic delays in CUIMC dataset compared to the claims databases. Nonetheless, more often than not, there are more diseases where DD for women exceeds that for men.

| | **Male anatomy conditions** | **Female anatomy conditions** | **Childbirth-related** |
|---|---|---|---|
| Remove if condition name string contains any of: | male, penis, testis, testicle | female, maternal, uterine, menopausal, uterus, umbilical, cervix, vaginal, vulva, placenta, ovary, postpartum, | pregnancy, gestation, birth, miscarriage, live born, obstet |

**Table S1. Anatomical keyword-list for removing sex-specific conditions in population-level characterization.** Sex-specific conditions that were removed from processing, as detailed in the population-level characterization methods from Supplementary Materials.

| Phenotype | Prec@50 | Prec@100 | Recall@50 | Rec@100* | F1@50 | F1@100 |
|---|---|---|---|---|---|---|
| **Crohn's** | **0.52** | 0.37 | 0.70 | **1.00** | **0.30** | 0.27 |
| **Gastritis** | **0.30** | 0.15 | 1.00 | **1.00** | **0.23** | 0.13 |
| **Hidradenitis** | **0.82** | 0.63 | 0.65 | **1.00** | 0.36 | **0.38** |
| **HIV** | **0.50** | 0.27 | 0.92 | **1.00** | **0.32** | 0.21 |
| **Osteoarthritis** | **0.80** | 0.50 | 0.80 | **1.00** | **0.40** | 0.33 |
| **Epilepsy** | **0.26** | 0.19 | 0.68 | **1.00** | **0.19** | 0.16 |
| **Migraine** | **0.18** | 0.14 | 0.64 | **1.00** | **0.14** | 0.12 |

**Table S2. Precision, recall, and F1-scores comparisons for the top-50 and top-100 algorithmically-generated relevant symptoms compared to clinician-adjudicated relevant symptoms.** Clinicians adjudicated a set of gold-standard relevant symptoms for a subset of chronic phenotypes, which we use assess our selection of the top-50 algorithmically-generated relevant symptoms for analysis. The top-50 algorithmically-generated symptoms consistently had the best trade-off between precision and recall compared to the top-100 symptoms. *Because clinicians adjudicated up to the top-100 symptoms and were asked to identify additional relevant symptoms (which none did), the recall@100 is always 1.00.

| Phenotype | Top-50 TTD diff (days) | Clinician TTD diff (days) |
|---|---|---|
| **Crohn's** | 30.43 | **40.04** |
| **Gastritis** | 69.66 | **126.03** |
| **Hidradenitis** | 50.19 | **172.86** |
| **HIV** | 0.69 | **14.56** |
| **Osteoarthritis** | **59.51** | 54.95 |
| **Epilepsy** | 72.26 | **161.72** |
| **Migraine** | 24.34 | **88.30** |

**Table S3. Comparison of time-to-diagnosis differences calculated using the top-50 algorithmically-generated relevant symptoms versus the clinician-adjudicated relevant symptoms using CCAE data.** The time-to-diagnosis (TTD) differences are consistently positive using both the algorithmically-generated and clinician-adjudicated relevant symptoms; in both cases, positive values indicate longer time-to-diagnoses for women. Clinical filtering essentially removed anomalous symptoms, and since the majority of symptoms are diagnosed later in women, removing anomalous symptoms kept the overall conclusions consistent, if not generally making the findings even more poignant.

| ICD-10-CM Chapter | Acute (1 year lookback) [31 phenotypes] | Mid-Length Chronic (3 year lookback) [65 phenotypes] | Long-Term Chronic (10 year lookback) [16 phenotypes] |
|---|---|---|---|
| I. Certain infectious and parasitic diseases | Sepsis, toxic shock syndrome, tuberculosis | Hepatitis C, Hepatitis Type B | Human Immunodeficiency Virus (HIV) |
| II. Neoplasms | Acute tubular necrosis | Chronic lymphoid leukemia, myelodysplastic syndrome, acute myeloid lymphoma, malignant tumor of bladder, malignant tumor of kidney, B-cell lymphoma, multiple myeloma, malignant tumor of stomach, non-Hodgkin's lymphoma, malignant tumor of colon, malignant neoplasm of liver, malignant neoplasm of respiratory tract | - |
| III. Diseases of the blood and blood-forming organs and certain disorders involving the immune system | - | Idiopathic thrombocytopenic purpura, thrombocytopenia | - |
| IV. Endocrine, nutritional, and metabolic diseases | - | Thyroiditis, Type 1 diabetes, hyperlipidemia, hypothyroidism | Type 2 diabetes |
| V. Mental, behavioral, and neurodevelopmental disorders | - | Schizophrenia, attention deficit hyperactive disorder, depression | Bipolar disorder, posttraumatic stress disorder, autism |
| VI. Diseases of the nervous system | Transient ischemic attack, acute disseminated encephalomyelitis, aseptic meningitis | Transverse myelitis, Multiple Sclerosis, Parkinson's, insomnia, hospitalization for Guillain-Barré | Sleep apnea, migraine incident, epilepsy, narcolepsy |
| VII. Diseases of the eye and adnexa | Optic neuritis | Glaucoma | - |
| VIII. Diseases of the ear and mastoid process | Otitis media | - | - |
| IX. Diseases of the circulatory system | Acute myocardial infarction, atrial fibrillation, heart failure, hemorrhagic stroke, stroke, pulmonary embolism, ischemic stroke, ventricular arrhythmia or cardiac arrest, deep vein thrombosis, cardiac arrhythmia | Thromboangiitis obliterans, myocarditis, hypertension, peripheral vascular disease, pulmonary hypertension | - |
| X. Diseases of the respiratory system | Pneumonia, respiratory failure, acute respiratory failure | Allergic rhinitis | - |
| XI. Diseases of the digestive system | Gastrointestinal bleeding, acute pancreatitis, hepatic failure | Constipation, cirrhosis, Celiac disease, Crohn's disease, gastroesophageal reflux | Ulcerative colitis, gastritis, inflammatory bowel disease |
| XII. Diseases of the skin and subcutaneous tissue | - | Psoriatic arthritis, erythema multiforme, atopic dermatitis and eczema | Hidradenitis suppurativa |
| XIII. Diseases of the musculoskeletal system and connective tissue | Thrombotic microangiopathy | Neck pain, osteoporosis, rheumatoid arthritis, sclerosis, lower back pain, Sjögren's, systemic lupus erythematosus, granulomatosis, multi-system inflammatory syndrome, ankylosing spondylitis, gout, Takayasu's disease, giant cell arteritis, vasculitis associated with ANCA | Osteoarthritis, Behçet's syndrome |
| XIV. Diseases of the genitourinary system | Acute kidney injury, kidney stone | Chronic kidney injury | - |
| XVIII. Symptoms, signs and abnormal clinical and laboratory findings, not elsewhere classified | Cardiogenic shock | Dysphagia, nausea, jaundice, abdominal pain, dyspnea, facial palsy, anosmia or hyposmia or dysgeusia | - |
| XIX. Injury, poisoning and certain other consequences of external causes | Chilblains, angioedema | - | - |

**Table S4. Phenotypes categorized by ICD-10-CM chapter.** The 112 phenotypes are split into 31 acute phenotypes (1 year lookback of presenting symptom), 65 mid-length chronic (3 years) phenotypes, and 16 long-term chronic (10 years of symptom data) phenotypes. The phenotypes cover all ICD-10-CM chapters except Chapter XV (Pregnancy, childbirth and the puerperium), Chapter XVI (Certain conditions originating in the perinatal period) and Chapter XVII (Congenital malformations, deformations and chromosomal abnormalities).

| Database & Phenotype Type | CCAE | | | MDCD | | |
|---|---|---|---|---|---|---|
| | **Acute** | **Chr. 3y** | **Chr. 10y** | **Acute** | **Chr. 3y** | **Chr. 10y** |
| % Symptoms Later | 86.3% | 83.2% | 69.6% | 60.8% | 70.3% | 71.0% |
| Avg. TTD diff (days) | 8.0 | 25.3 | 51.9 | 2.0 | 12.9 | 53.6 |
| Nb sig. DD | 31 | 62 | 15 | 28 | 56 | 14 |
| Sig. DD diff (days) | 21.0 | 62.9 | 134.0 | 11.4 | 42.3 | 89.7 |

| Database & Phenotype Type | MDCR | | | CUIMC | | |
|---|---|---|---|---|---|---|
| | **Acute** | **Chr. 3y** | **Chr. 10y** | **Acute** | **Chr. 3y** | **Chr. 10y** |
| % Symptoms Later | 68.2% | 74.7% | 72.6% | 55.5% | 54.0% | 60.4% |
| Avg. TTD diff (days) | 2.4 | 13.9 | 69.0 | -1.4 | 5.5 | 78.2 |
| Nb. Sig. DD | 22 | 47 | 10 | 9 | 27 | 12 |
| Sig. DD diff (days) | 5.1 | 21.9 | 21.3 | 3.3 | 17.6 | 211.1 |

**Table S5. Summary statistics for delays in diagnosis between women and men.** Percentage of symptoms with delayed diagnosis in women; average time-to-diagnosis (TTD) differences across the top-50 algorithmically-generated presenting symptoms (where positive values indicate a longer TTD for women); number of significant diagnostic delay phenotypes (out of 31 for acute, 65 for chronic 3 years, and 16 for chronic 10 years); and average diagnostic delay difference in days for the significant phenotypes (where positive values indicate a longer DD for women).
